# Modeling and analysis of COVID-19 infected persons during repeated waves in Japan

**DOI:** 10.1101/2021.10.11.21264869

**Authors:** Koichi Hashiguchi

**Affiliations:** None

## Abstract

A model for estimating the number of COVID-19 infected persons (infecteds) is proposed based on the exponential function law of the SIR model. This model is composed of several equations expressing the number of infecteds, considering the onset after an incubation period, infectivity, wavy infection persistence with repeated infection spread and convergence with insufficient quarantine. This model is applied to the infection in Japan, which is currently suffering from the 5th wave, and the initial number of infecteds and various related parameters are calculated by curve fitting of the cumulative number of infecteds to the total cases in the database. As a minimum boundary of the number of infecteds for the infection continuation up to the 5th wave, the initial number of infecteds at the outbreak of infection is more than an order of magnitude higher than the actual initial cases. A convergence ratio (cumulative number of infecteds / total cases) at the end of the first wave is obtained as 1.5. The quarantine rate and social distancing ratio based on the SIQR model are evaluated, and the social distancing ratio increases sharply just after the government’s declaration of emergency. The quarantine rate closely equals the positive rate by PCR tests, meaning that the number of infecteds, which had been unknown, is on the order of almost the same as the number of tests.

## Introduction

A new type of coronavirus, COVID-19, which took off in Wuhan, China, at the end of 2019 and quickly spread all over the world, has not been settled even after one year and a half, with repeated waves of spread and convergence of the infection.

The SIR model ^1)^ proposed about 100 years ago is known as a mathematical model for describing the epidemic process of infection. It is a simple model that expresses the increase/decrease in the number of persons between groups S (Susceptible), I (Infected), and R (Recovered) with differential equations under the population conservation law. This model reproduced the plague epidemic of the early 20th century and is considered to help understand the current COVID-19 epidemic ^2).^ Furthermore, an improved SIQR model ^3)^ was also proposed, in which the recovered group, R in the SIR model was divided into Q (quarantined), and R (recovered). Odagaki ^4)^ proposed SIQR model with two important parameters, the quarantine rate and the social distancing ratio. These explicitly introduced parameters are related with the exponent λ of an exponential function for the daily variation in the number of new infection-positive persons and contribute to the evaluation of infection control measures.

In the previous papers ^5, 6)^ intended to analyze these parameters, mathematical analyses between these relationships were conducted on Taiwan and South Korea with measures of mass PCR tests in the early stages of infection, and Western countries with measures of lockdown, including Japan, and clarified the difference in the quarantine rate-social distancing ratio relationship caused by these countermeasures.

In this paper, a model estimating the number of infected persons, which has been unknown in the previous papers, is proposed with consideration of an incubation period after infection, subsequent infection rate, and the convergence ratio at the stage of wavy infection convergence. This model also presupposes the exponential spread of infection common to the SIR and SIQR models under the law of conservation of population. This model is applied to a database of infection status, and the daily variation of the infected persons is evaluated by fitting the calculated cumulative number of infected persons to the actual value of the cumulative number of newly detected infection-positive persons. Using the results, the variation of infected persons, quarantine rate, and social distancing ratio are analyzed during five waves in Japan.

### Basic equations in the model

Basic equations in this estimation model are described separately for the derivation of the cumulative number of infected persons, and the derivation of the quarantine rate / social distancing ratio. Since the latter parameters have been described in detail in the previous papers, only the resulting equations are described.

### Basic equations for the cumulative number of infected persons

Assuming the cumulative number of infected persons (hereinafter referred to as cumulative infecteds) *Z*_i_ on *d*_i_ day increases exponentially, *Z*_i_ is expressed by the exponential function (1) using an exponent *α*_i_.

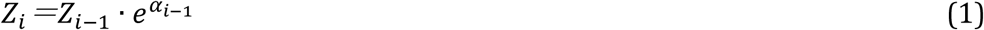

The number of newly infected persons Δ*Z*_i_ is expressed by Eq. (2) as the increase in cumulative infecteds.

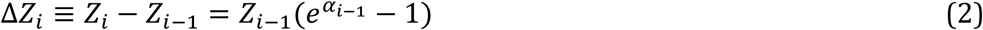

After infection, a latent disease declares itself after an incubation period. Assuming an incubation period follows the gamma distribution *f(d)* with maximum onset on the 5th day after infection and ending on the 16th day ^7)^, which gives the shape parameter a = 5 and the scale parameter b = 1.25 in the gamma distribution formula. This gamma distribution curve is discretized, normalized so that the sum of *f(d)* for 16 days is 1, and the ratio of onset on day *d*_j_ is expressed by a function *gm*_j_ in Eq. (3).

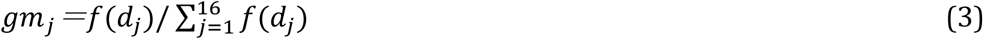

The number of infected persons Δ*Z*_*i*_ on day *d*_*i*_ is ivided among 16 days *d*_*i*+1_ to *d*_*i*+16_ according to the gamma distribution, the number of onsets on day *d*_*j*_ is Δ*Z*_*i*_*·gm*_*j*_. Therefore, the number of onsets *S*_*i*_ on day *d*_*i*_ is the sum of the number of persons who have been infected from 16 days before and have completed the incubation period, Eq. (4) is obtained.

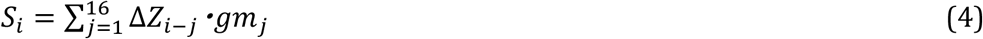

The number of infected persons (hereinafter referred to as infecteds) *X*_*i*_ on *d*_*i*_ day is expressed in Eq. (5), as the sum of infecteds *X*_*i*-*1*_ on a previous day and the number of cases *S*_*i*_ that completed the incubation period with subtracting the number of newly detected and quarantined persons *y*_*i*_ (new cases).

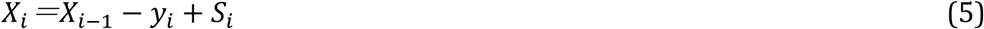

The cumulative infecteds *Z*_*i*_ is given by Eq. (6).

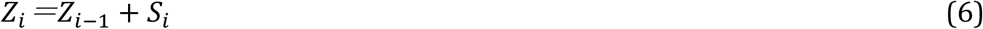

In these basic equations, setting the initial infecteds at the outbreak of infection as *X*_0_, which is equal to the cumulative infecteds *Z*_0_ on the infection outbreak, and this initial value is substituted into Eq. (2). By repeating the sequential calculation from equations (4) to (6), the infecteds and the cumulative infecteds during a predetermined period are obtained. The basic equations mentioned above are all linear form, and *Z*_*i*_ in Eq. (6) increases and decreases linearly corresponding to the increase and decrease in the initial value *Z*_0_. The calculated cumulative infecteds are analyzed by referring to the transition data of the cumulative number of new cases (total cases *z*_*i*_) in the database. In these basic expressions, uppercase variables such as *Z*_*i*_,*X*_*i*_,*S*_*i*_ are expressed as calculated values related to infecteds, and lowercase variables such as *y*_*i*_,*z*_*i*_ are expressed as actual values related to cases from the database.

Infection progresses in a wavy way in which the number of new cases repeatedly increases and decreases, and in the calculation process for each wave, two parameters, *IR* (Infection Ratio) and *CR* (Convergence Ratio), are introduced to ensure consistency between waves. The former is defined as the proportion of infecteds capable to infect others after the incubation period, and Δ*Z*_*i*_ in Eq. (2) is replaced by Eq. (2’).

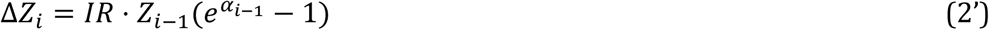

The latter *CR* is defined as a ratio of the calculated cumulative infecteds *Z*_*i*_ to the total cases *z*_*i*_ at the end of each wave. When *CR* = 1, excess infecteds becomes 0, which means all the cumulative infecteds are detected and quarantined as total cases during the period, resulting in termination of infection. Otherwise, excess infecteds with *CR* larger than 1 bring about the next spread of infection. By definition, both parameters can be varied within a range of *IR* = 0 *∼* 1’*CR* ≥ 1.

### Basic equations for quarantine rate and social distancing ratio

The new cases *y*_*i*_ is expressed by Eq. (7) using an exponent λ.

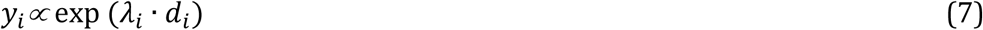

As already mentioned in the previous papers, the quarantine rate *QR* and the social distancing ratio *SD* (q and x, respectively in the previous papers) are shown by equations (8) and (9), respectively. The relationship of these parameters and equations in Eq. (7) is expressed by the following equations.

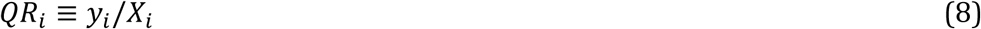

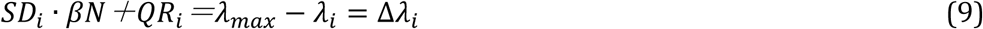

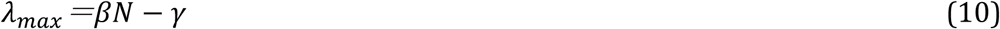

Both parameters are constrained in the range of 0 ≤ *SD*_*i*_, *QR*_*i*_ ≤ 1 by definition. *βN* is the transmission coefficient *β* multiplied by population N, which is an index of the degree of transmission due to contact between uninfected and infected persons. *γ* is the cure rate calculated as 0.04 reciprocal of the healing period set to 25 days ^2)^. However, *γ* is set as *−λ*_*min*_, which is obtained by substituting Eq. (9) into the constraint of 0 ≤ *SD*_*i*_ ≤ 1, because 0.04 for *γ* may give results deviating from upper constraint for *SD*. The unit of λ is (1 / day), *QR* and *βN* have the same units, and *SD* is dimensionless. *βN* associated with the maximum λ in Eq. (10) is regarded as invariant throughout the infection period.

### Analysis of COVID-19 infecteds in Japan

This model was applied for the analysis of the infection status in Japan. Daily variation of new cases, *y*_*i*_ and total cases, *z*_*i*_ from the “Our World in Data” (January 1, 2020-August 4, 2021) database ^8)^ were converted to per million population, as well as the number of PCR tests described later. As a preliminary preparation, the daily *α*_*i*_ and *λ*_*i*_ were calculated by the logarithmic regression of Eq. (1) and (7) using those converted data. In the regression calculation, the exponent was calculated as the moving-average logarithmic slope for 13 days, and *gm*_*j*_ of Eq. (3) was also calculated. The variation of new cases, total cases, and the corresponding exponents λ and α are shown in Fig. 1a, and b. In Fig. 1a, the new cases repeatedly increase and decrease exhibiting four waves, and currently is rapidly increasing as the fifth wave. In the figure, the converging points with minimum new cases between peakss are indicated by red dots. The total cases exhibit a gentle stepwise increase near the converging points with a rapid increase at the maximum points of new cases in each wave. The variation of exponents *α*_*i*_ and *λ*_*i*_ shown in Fig. 1b are mathematically correlated as differential and integral calculus. The major difference is that λ can be positive and negative, however, α is always 0 or positive.

**Fig. 1.**
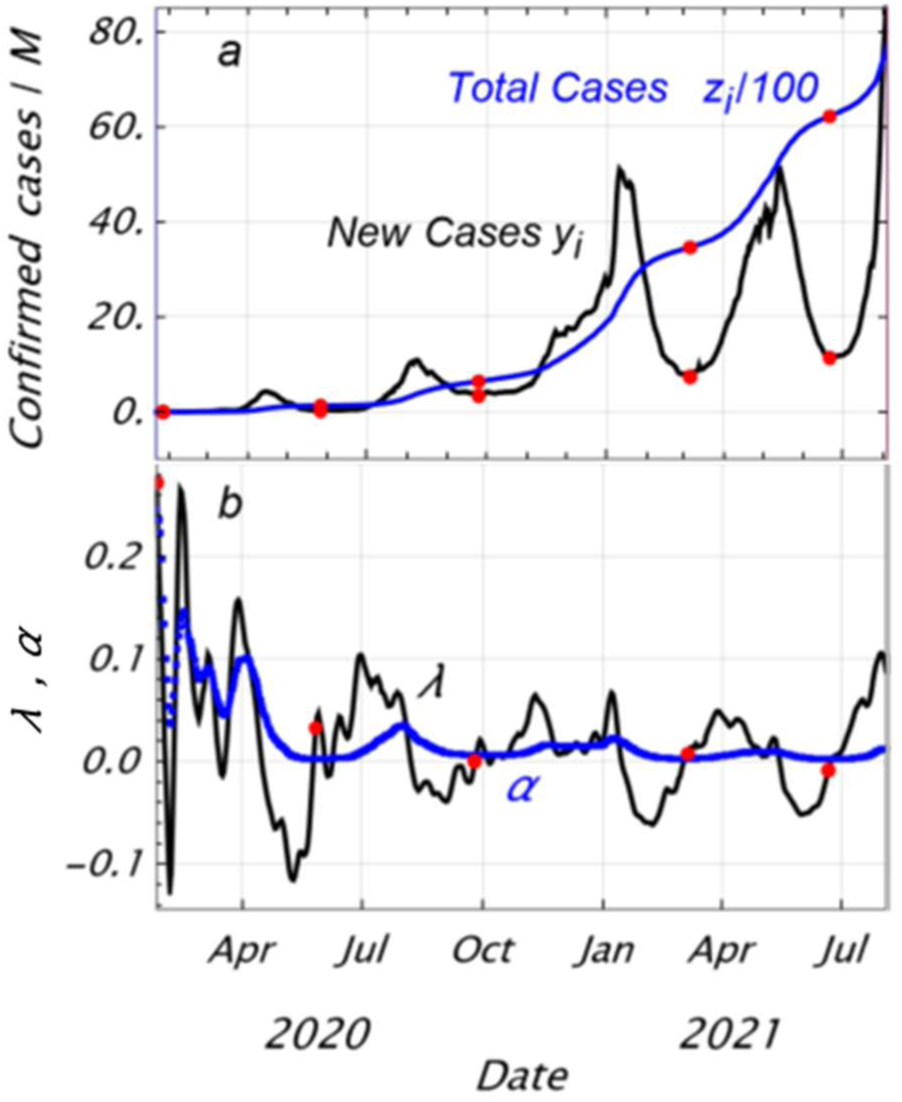
Variation of new cases, total cases plotted in scale 1/100 (a) and corresponding exponents of λ and α (b) of COVID-19 infection in Japan,data from “Our World in Data”. Red dots showing converging points for each wave.

Using the above basic equations, the cumulative infecteds *Z*_*i*_ is calculated sequentially day by day for a given initial value *X*_0_(= *Z*_0_) at the outbreak of infection. Any calculated *Z*_*i*_ curve above the transition *z*_*i*_ curve in Fig. 1a is acceptable, however, the procedure for obtaining the optimum solution is performed by finding the lower limit for fitting the calculated value *Z*_*i*_ to the actual value *z*_*i*_ transition data. In the following calculation, the infection ratio *IR*_n_ and convergence ratio *CR*_n_ were kept constant in the same wave and varied for each wave n (n = 1 to 4). Since *Z*_*i*_ = *CR*_n_ · *z*_*i*_ is set at a converging point of each wave, increase in *CR*_n_ increases *Z*_*i*_ at the converging point, while decrease in *IR*_n_ decreases *Z*_*i*_ by Eq. (2’). Therefore, the increase/decrease in both parameters has an adjusting function of *Z*_*i*_ between waves at the converging point of each wave. Since there was no termination of infection during this calculation period, cumulative infecteds is always greater than or equal to the actual value (*Z*_*i*_ ≥ *z*_*i*_), and infecteds is restricted to be greater than or equal to the new cases (*X*_*i*_ ≥ *y*_*i*_).

#### First wave

The outbreak day of infection is set on January 27, 2020, as a day of *λ*_*max*_ = 0.272, and an arbitrary value is set for the initial value *X*_0_ (=*Z*_0_) on this day. The daily variation of *Z*_*i*_ are sequentially calculated using *α*_*i*_ and *y*_*i*_ in Fig. 1b based on Eq. (2’) to Eq. (6) via daily Δ*Z*_*i*_, *S*_*i*_, *X*_*i*_ to the converging point of the first wave. The convergence calculation is applied to find *X*_0_, which minimizes the difference between *Z*_*i*_ and *CR*_1_ · *z*_*i*_ at the converging point of 1st wave. On this calculation, *CR*_1_ value was varied in 0.1 step within the range of 1 to 2.3 and the calculation was executed to obtain *X*_0_ for each *CR*_1_ value. In the 1st wave, all infected persons are assumed to be infectivity, then *IR*_1_ was set to be 1.

#### After the 1st wave

The same calculating procedure is adopted for subsequent waves setting the last *X*_*i*_ value of previous wave as the initial value for the next wave. For each *CR*_1_ value of the first wave, the infection ratio *IR*_n_ (n = 2 *∼* 5) is varied in a range of 0.1*∼*1 by 0.01 step in the new wave. Among those calculations, the value *IR*_n_ minimizing *Z*_*i*_ *− z*_*i*_ (> 0) at the converging point of each wave is chosen as the best fit and *CR*_n_ is calculated as *CR*_n_ = *Z*_*i*_/*z*_*i*_. This process is repeated for the subsequent waves. There is some constraint for the allowable range of the initial value of infecteds *X*_0_ in the first wave due to the linearity between *X*_0_ *and Z*_*i*_ as shown in Fig. 2. A higher value of *X*_0_ gives higher *X*_*i*_, which is acceptable. On the other hand, lower *X*_0_ gives lower *X*_*i*_, then the term of the new cases *y*_*i*_ in Eq. (5) prevails compared to *X*_*i*-1_ and *S*_*i*_, resulting in a large and rapid decrease in *X*_*i*_ into negative value.

**Fig. 2.**
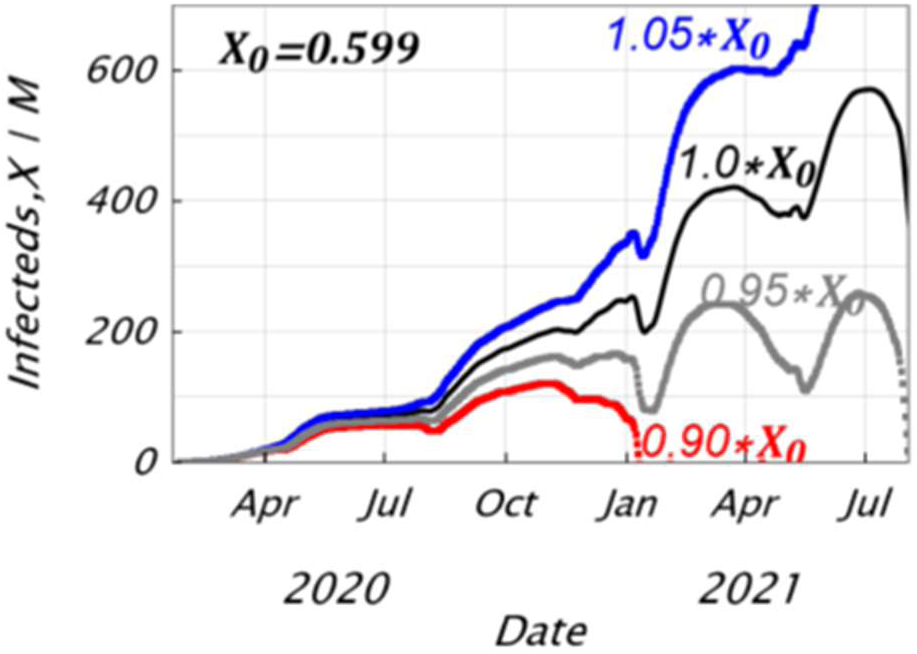
Effects of initial value of infected persons at outbreak on variation in number of infected persons (infecteds). Lower initial value leads to early termination.

Based on these considerations, the calculation up to the 5th wave was carried out with the condition that the cumulative infecteds *Z*_*i*_ curve up to the final wave best fits the total cases *z*_*i*_ curve. This gives the lower limit of *Z*_*i*_. As a condition for the best fit, an index that minimizes the difference between *Z*_*i*_ and *z*_*i*_ over the entire period expressed by Eq. (11) is introduced.

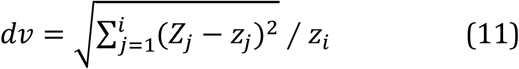

The relationship between *dv* (%) in Eq. (11) and *CM*_1_ value in the first wave is shown in Fig. 3a. *dv* values obtained are all little over 0.1%, showing very good fittability. As shown in Fig. 3b, the initial value *X*_0_ of infecteds increases linearly with the increase in the convergence ratio *CR*_1_ of the first wave. In the range of *CR*_1_ <1.5, *X*_0_ becomes lower and *X*_*i*_ decreases sharply as shown in Fig.2 before reaching the 5th wave. Thus *X*_0_ is determined as a condition necessary for the continuation of infection up to the fifth wave. In addition, for the purpose of searching for the lower limit of the cumulative infecteds and reducing the number of calculations, the analyses were limited in a range of *dv* lower than 0.15.

**Fig. 3.**
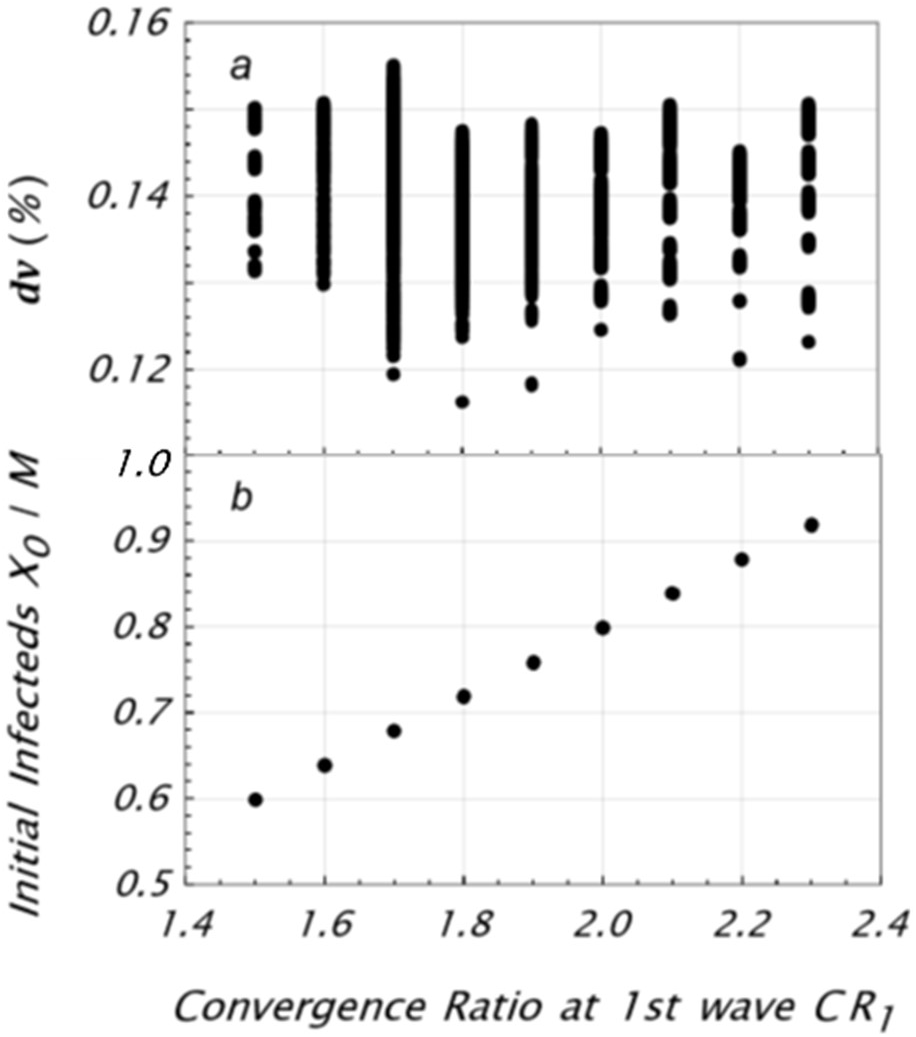
Relationship between 1st wave convergence ratio, *CR*_1_ and best fit index, dv over the entire period (a), and initial value of infecteds, *X*_0_ (b). Initial value varies linearly with convergence ratio.

The variation of infecteds *X*_*i*_ and cumulative infecteds *Z*_*i*_ are shown in Fig. 4a, b for each *CR*_1_ value of 1.5, 1.9, and 2.3 with minimum *dv*. In the case of a lower convergence ratio *CR*_1_ of the first wave, infecteds *X*_*i*_ stay lower in the early wave and higher in the latter wave compared to the higher *CR*_1_ case. The cumulative infecteds in Fig. 4b exhibits the best fittability with the small deviation from the total cases in the latter wave. Table 1 shows the *CR*_n_ and *IR*_n_ values for each wave including conditions other than those in Fig.4. An increase in *CM*_1_ results in a decrease in *IR*_2_, and subsequent convergence ratios remains close to 1.

**Fig. 4.**
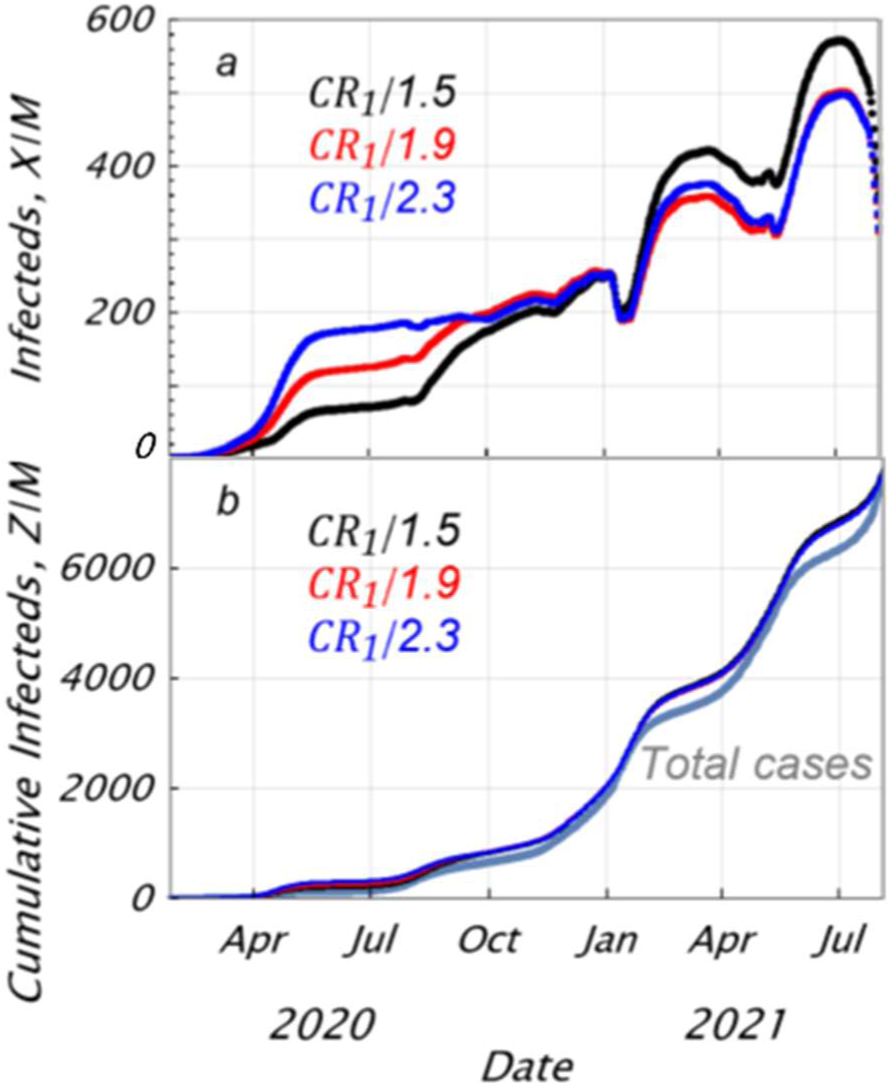
Variation of infecteds *X*_*i*_ (a) and cumulative infecteds *Z*_*i*_ (b) for each *CR*_1_ value of 1.5, 1.9, and 2.3. Cumulative infecteds best fit to total cases.

**Table 1.**
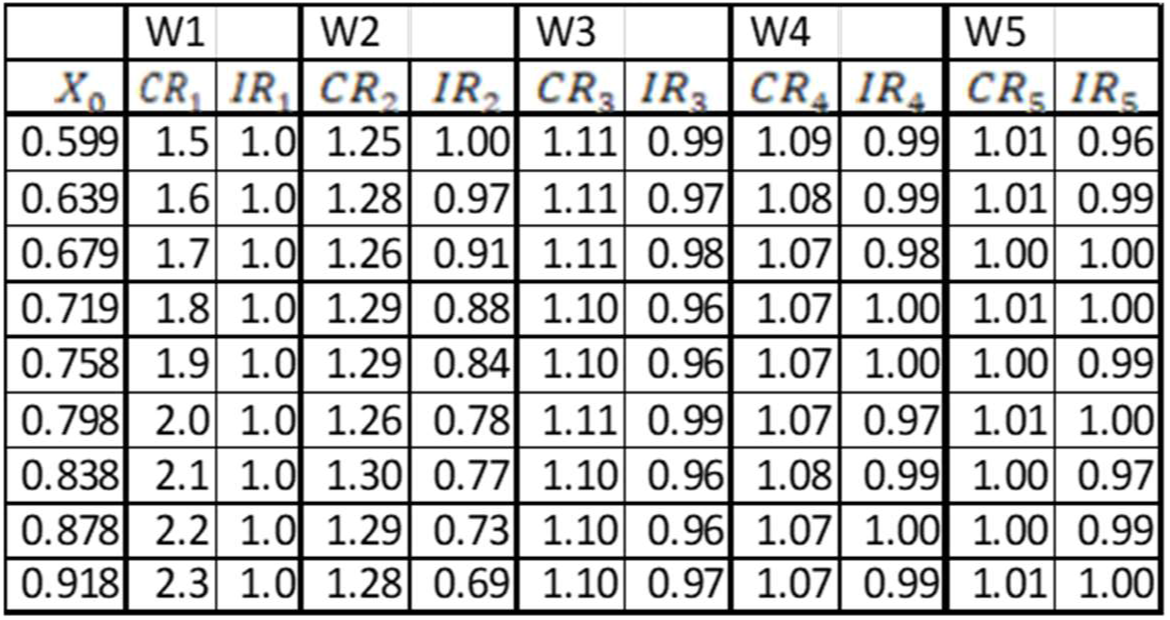
*CR*_*n*_ value, *IR*_*n*_ value in each wave

|  | W1 |  | W2 |  | W3 |  | W4 |  | W5 |  |
| --- | --- | --- | --- | --- | --- | --- | --- | --- | --- | --- |
| $X_0$ | $CR_1$ | $IR_1$ | $CR_2$ | $IR_2$ | $CR_3$ | $IR_3$ | $CR_4$ | $IR_4$ | $CR_5$ | $IR_5$ |
| 0.599 | 1.5 | 1.0 | 1.25 | 1.00 | 1.11 | 0.99 | 1.09 | 0.99 | 1.01 | 0.96 |
| 0.639 | 1.6 | 1.0 | 1.28 | 0.97 | 1.11 | 0.97 | 1.08 | 0.99 | 1.01 | 0.99 |
| 0.679 | 1.7 | 1.0 | 1.26 | 0.91 | 1.11 | 0.98 | 1.07 | 0.98 | 1.00 | 1.00 |
| 0.719 | 1.8 | 1.0 | 1.29 | 0.88 | 1.10 | 0.96 | 1.07 | 1.00 | 1.01 | 1.00 |
| 0.758 | 1.9 | 1.0 | 1.29 | 0.84 | 1.10 | 0.96 | 1.07 | 1.00 | 1.00 | 0.99 |
| 0.798 | 2.0 | 1.0 | 1.26 | 0.78 | 1.11 | 0.99 | 1.07 | 0.97 | 1.01 | 1.00 |
| 0.838 | 2.1 | 1.0 | 1.30 | 0.77 | 1.10 | 0.96 | 1.08 | 0.99 | 1.00 | 0.97 |
| 0.878 | 2.2 | 1.0 | 1.29 | 0.73 | 1.10 | 0.96 | 1.07 | 1.00 | 1.00 | 0.99 |
| 0.918 | 2.3 | 1.0 | 1.28 | 0.69 | 1.10 | 0.97 | 1.07 | 0.99 | 1.01 | 1.00 |

For the case of *CR*_1_ value 1.5, the quarantine rate *QR*_*i*_ and the social distancing ratio *SD*_*i*_ are calculated using the equations (8) and (9) and shown in Fig. 5a, b. By definition, peaks of quarantine rate correspond to the peak of new cases in each wave and reach up to about 0.25 in the third wave, staying low in other waves. Higher *CR*_1_ makes *QR*_*i*_ increase in the later wave due to decrease in *X*_*i*_ in later wave.

**Fig. 5.**
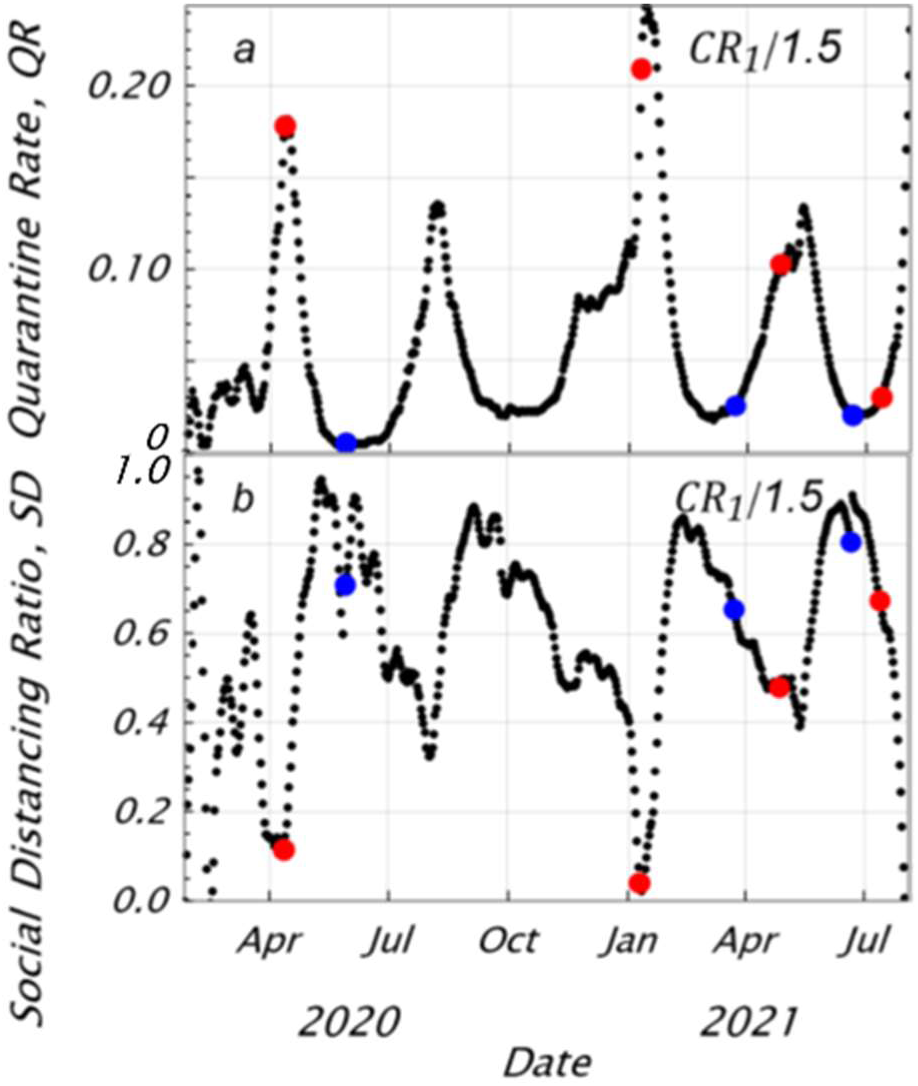
Transition of quarantine rate *QR* (a) and social distancing ratio *SD* (b), red dots: declaration of emergency, blue dots: cancellation of emergency

The social distancing ratio exhibits the opposite tendency to the quarantine rate and tends to be maximum near the minimum quarantine rate point. This is because the sum of the quarantine rate *QR*_*i*_ and the social distancing ratio *SD* equals Δ*λ*_*i*_ in Eq. (9), and the variation in Δ*λ*_*i*_ is small in a short period. In this figure, the dates of declaration and cancellation of the government’s state of emergency is indicated by red and blue dots, respectively. In each wave, the social distancing ratio increases sharply on the declaration, peaks with a time lag of about one month, and then the declaration is lifted after the small decrease in social distancing ratio. The social distancing ratio at the first declaration reached nearly 1, and this high level lasted for a long period, indicating a greater declaration effect, however, the peaks after the second wave become a little bit lower and shorter than the first wave, showing weakening declarative effect.

The loci of the relationship between the quarantine rate and the social distancing ratio are shown in Fig. 6 separately for each wave. In each wave, the social distancing ratio decreases as the quarantine rate increases during infection spread, and vice versa at the stage of infection convergence. These loci show a tendency similar to the loci analyzed in the previous paper.

**Fig. 6.**
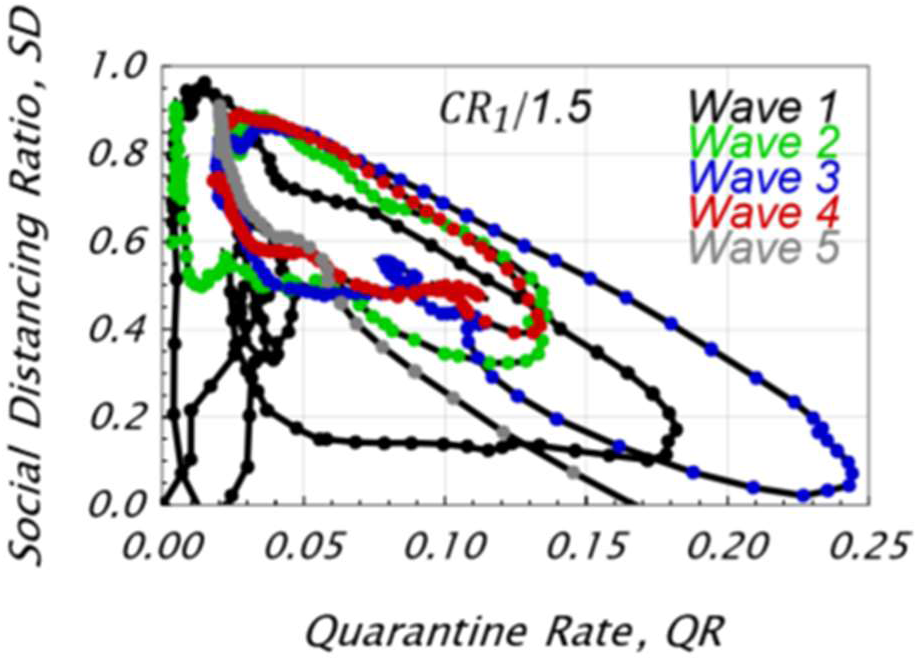
Relationship between quarantine rate and social distancing ratio for each wave

The variation of positive rate by PCR test as the measures of quarantine is shown in Fig. 7a together with the variation of quarantine rate. The positive rate varies at almost the same timing and same level as the quarantine rate. Both numerators have the same number of new cases, which means that both denominators, the number of tests and the infecteds *X*_*i*_ might be nearly equal. As shown in Fig. 7b, the number of PCR tests and the number of infecteds *X*_*i*_ vary at almost the same level. This means that the number of tests is considerably insufficient for detecting and quarantining newly infected persons. Fig. 8 shows the effects of varying the quarantine rate in Fig. 5a between 0.95 and 1.1 times on the transition of the infecteds by simulation. An increase in the quarantine rate decreases the infecteds, and 10% increase in the quarantine rate may terminate the infection after the third wave. Increasing the quarantine rate also reduces the exponent α together with *CR*_1_, which may further accelerate the termination of infection.

**Fig. 7.**
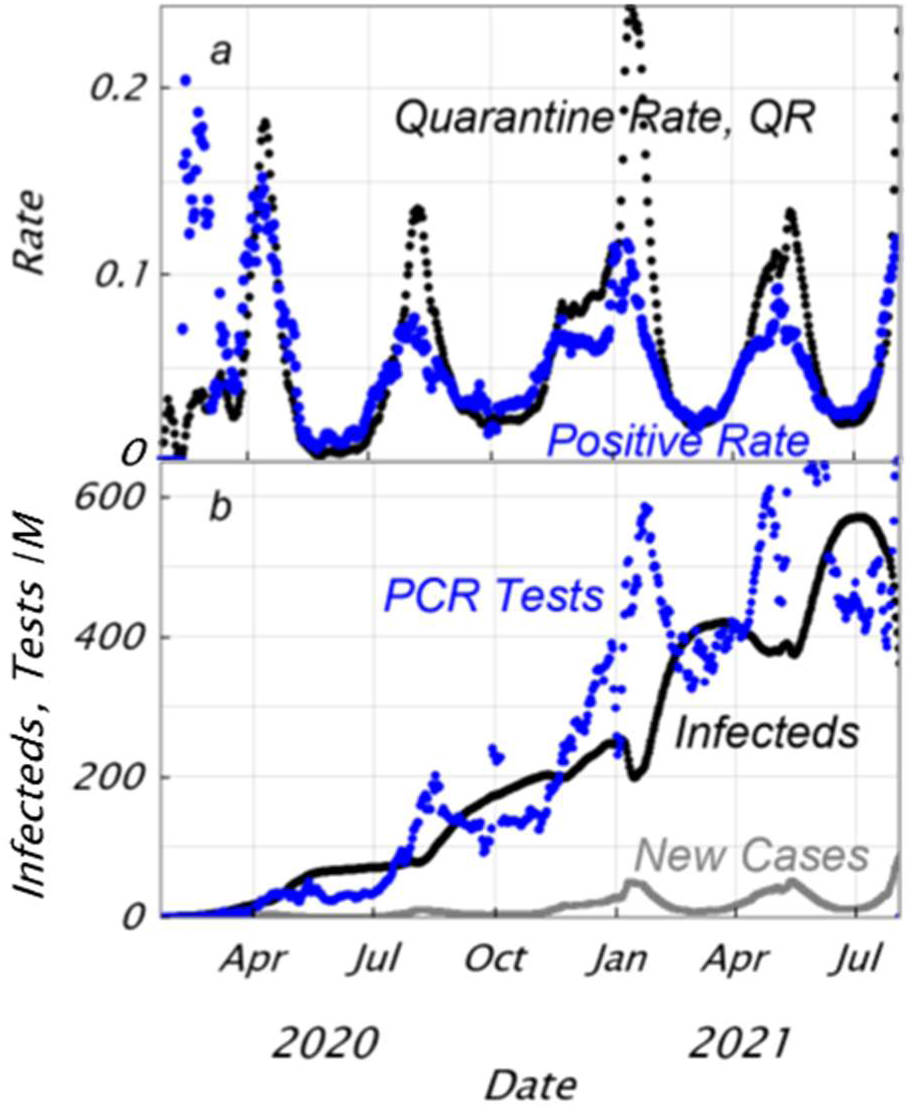
Variation of positive rate in PCR tests and quarantine rate (a), and comparison of PCR tests and infecteds(b); QR and positive rate are varying in the same level.

**Fig. 8.**
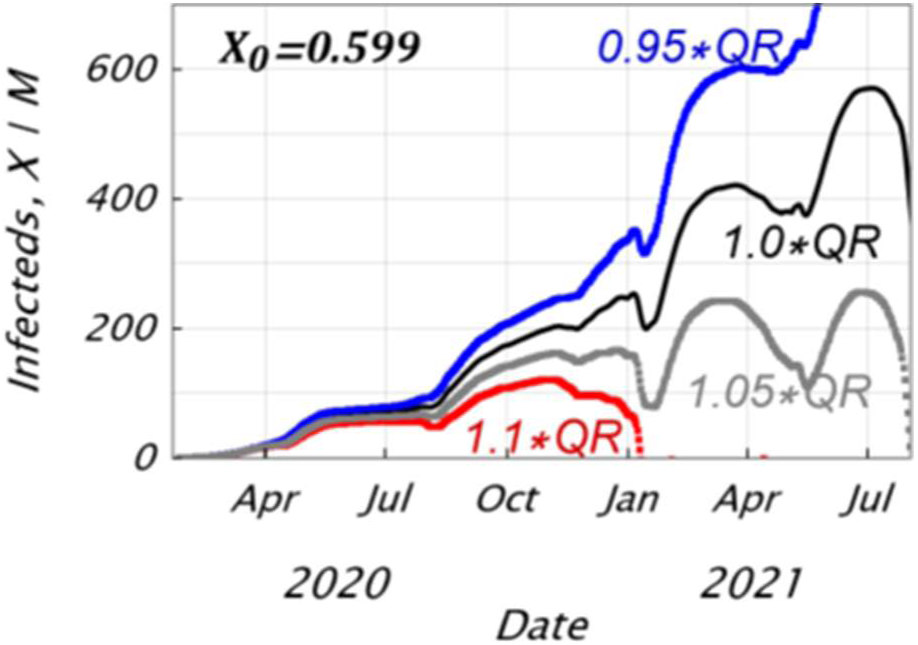
Effect of quarantine rate, QR on variation of infecteds; increase in QR accelerates convergence of infection.

The calculation for the best fit of the cumulative infecteds were executed for a lot of combinations of parameters, *X*_0_, *CR*_1_ for the 1st wave, and *IR*_n_ for each wave. Apart from this method, there is also a deductive way, using the linearity between *X*_0_ and *X*_*i*_ or *Z*_*i*_ as shown in Fig. 3b. Relational expressions between the initial value *X*_0_ and *Z*_*i*_ are formulated for each *IR*_n_ for each wave, and the optimum solution is obtained while matching the waves using these relational expressions. However, it is necessary to pay attention to the case where the incubation period expressed by Eq. (3) spans two waves.

## Conclusions

A model for estimating the number of COVID-19 infected persons (infecteds) was proposed based on the exponential function law of the SIR model. This model is composed of several equations expressing infecteds in the infection process, considering the onset after an incubation period, infectivity, wavy infection persistence with repeated infection spread and convergence while insufficient quarantine. Furthermore, the model was applied to each wave, where parameters were introduced to guarantee the continuity and compatibility between neighboring waves. This model was applied to the infection in Japan, which is currently up to the 5th wave, and the initial infecteds and various parameters were calculated by curve fitting of the cumulative infecteds to the total cases in the database.

- As a best-fit solution of the cumulative infecteds up to the 5th wave to the total cases, the initial infecteds of 0.6 / million population at the start of infection is more than an order of magnitude higher than the actual initial cases of 0.03 / million population. A convergence ratio 1.5 (cumulative infecteds / total cases) at the end of the first wave was obtained.
- This result gives a minimum value of infecteds for the infection continuation up to the 5th wave. Otherwise, the initial infecteds less than this lower limit may result in the termination of infection in the second or the third wave.
- Although a convergence ratio of more than 1.5 in the first wave is allowed, the number of excess infecteds, in that case, is compensated by the decrease in the infection ratio (infectivity ratio of infecteds after the incubation period) in the second wave.
- The isolation rate and social distancing ratio based on the SIQR model proposed by Odagaki were evaluated, and the social distancing ratio increases sharply just after the government’s declaration of emergency. The quarantine rate closely equals the positive rate by PCR tests, meaning that the infecteds, which had been unknown, is the same level as the number of tests.

## Data Availability

All data produced are available online at Our World in Data COVID-19 dataset

https://ourworldindata.org/coronavirus-source-data

## Acknowledgment

The author would like to thank Professor Emeritus Ikuo Yoshihara of the University of Miyazaki for his detailed suggestions and discussions.

